# Enteral docosahexaenoic and arachidonic acid supplementation and retinopathy of prematurity: a re-analysis of randomized controlled trials in preterm infants

**DOI:** 10.64898/2026.06.12.26355524

**Authors:** Ulrika Sjöbom, Aldina Pivodic, Pia Lundgren, Sissel J Moltu, Brandy Frost, Daniel T Robinson, Christine Henriksen, Ann Hellström, Anders K Nilsson

## Abstract

**Background:** A recent meta-analysis by Dang *et al. [1]* concluded that enteral supplementation with docosahexaenoic acid (DHA), with or without arachidonic acid (ARA) did not significantly affect retinopathy of prematurity (ROP) outcomes in preterm infants. Of four eligible trials that supplemented both DHA and ARA, only two contributed to each ROP outcome analyzed, and severe ROP was not assessed.

**Methods:** We replicated the eligibility criteria and search strategy of Dang *et al*., restricted to trials that supplemented both DHA and ARA, and reanalyzed three ROP endpoints (any ROP, ROP requiring treatment, and severe ROP [stage 3 and/or treated]) using complete outcome records from all eligible trials. Crude risk ratios (RR) were pooled by Mantel-Haenszel fixed-effect meta-analysis. Gestational age-adjusted odds ratios (adjOR) were pooled on the log scale by inverse-variance random-effects meta-analysis with restricted maximum likelihood (REML) estimation of between-study variance and Hartung-Knapp confidence intervals.

**Results:** Five trials were included; one trial was identified in our replicated search but was excluded by Dang *et al*. without a stated rationale. The pooled estimate for any ROP was consistent with Dang *et al*. (RR 0.87 [95% CI 0.71–1.08]; adjOR 0.70 [0.46–1.08]). For ROP requiring treatment, the crude RR suggested a lower risk but did not reach statistical significance (RR 0.60 [0.35–1.04]), whereas the gestational age-adjusted estimate indicated lower odds (adjOR 0.47 [0.23–0.94]). For severe ROP, DHA+ARA supplementation produced a significant protective effect in both unadjusted and adjusted models (RR 0.56 [0.36–0.86]; adjOR 0.42 [0.19–0.96]).

**Conclusions:** When all eligible trials contribute to each endpoint and severe ROP is included as an outcome, enteral DHA+ARA supplementation reduces severe ROP and is associated with lower odds of ROP requiring treatment after adjustment for gestational age. These findings differ from the conclusions of Dang *et al*. and support reconsideration of DHA+ARA supplementation as a strategy to reduce sight-threatening ROP in preterm infants.

## Introduction

Retinopathy of prematurity (ROP) is a vasoproliferative retinal disorder affecting preterm infants. Severe ROP, typically defined as stage 3 disease or disease requiring treatment, carries a risk of lifelong visual impairment, including blindness [2]. Several modifiable risk factors have been identified, including oxygen exposure and nutritional status, and nutritional interventions to reduce the risk of severe ROP are an active area of research [3-5].

Long-chain polyunsaturated fatty acids (LCPUFAs), notably docosahexaenoic acid (DHA, 22:6n-3) and arachidonic acid (ARA, 20:4n-6), are essential structural and signaling lipids that accumulate in the retina and central nervous system during the third trimester. Preterm birth interrupts this accumulation, and exogenous postnatal supply via formula or fortified human milk does not necessarily restore adequate levels [6-8]. In murine models, oral DHA with ARA reduced vaso-obliteration in oxygen-induced retinopathy and promoted physiological retinal vessel growth in hyperglycemia-induced retinopathy [9] Maintaining a balance between ω-3 and ω-6 LCPUFAs has been shown to be required for normal retinal vascular and neuronal development in the same Phase I ROP model [10]. However, randomized controlled trials of enteral DHA, with or without ARA, in preterm infants have produced inconsistent results for ROP [11].

Dang *et al*. [1] recently published a systematic review and meta-analysis of the effect of enteral DHA supplementation, with or without ARA, on neonatal morbidities, including ROP. They identified four randomized controlled trials that supplemented preterm infants with both DHA and ARA. For the ROP analyses, however, only two trials contributed to “any ROP” [12, 13] and only two to “ROP requiring treatment” [14, 15], leaving half of the eligible evidence outside each pooled estimate. The authors reported no statistically significant effect of DHA+ARA on ROP outcomes and concluded that enteral supplementation did not confer protective effects against major complications of prematurity.

We hypothesized that this null conclusion for ROP was driven by incomplete inclusion of available outcome data rather than by a true null effect. We therefore re-analyzed the same body of evidence using complete records for each ROP outcome from all eligible trials supplementing both DHA and ARA, additionally included the more clinically relevant outcome of severe ROP, and reported gestational age–adjusted odds ratios alongside crude risk ratios.

## Methods

### Eligibility criteria and search strategy

We followed the eligibility criteria and search strategy specified in the protocol used by Dang *et al*. (PROSPERO CRD42024552578) [16], and restricted the search to trials supplementing both DHA and ARA. In brief, randomized controlled trials of enteral DHA with ARA, initiated within 28 days of birth compared with placebo or no intervention, in preterm infants (primarily gestational age <34 weeks or birth weight <2000 g) were eligible. No restrictions were placed on dose or duration of supplementation.

### Included studies and data extraction

Five trials were included: Hellström *et al*. 2021 [12], Moltu *et al*. 2023 [13], Frost *et al*. 2021 [14], Robinson *et al*. 2016 [15], and Henriksen *et al*. 2008 [17]. The first four were identified and used by Dang *et al*.; the fifth, Henriksen *et al*. 2008 [17], was identified by their search strategy but excluded from their analysis without a stated rationale. Outcome data (the number of infants per arm and the number with any ROP, severe ROP, and ROP requiring treatment) were obtained directly from the original trial datasets.

### Risk of bias

Risk-of-bias assessments for the four trials included in the Dang *et al*. meta-analysis were taken from their published assessment. Because the Henriksen *et al*. trial was not included by Dang *et al*., no risk-of-bias rating was available from their work, and we therefore performed an independent assessment for that trial using the Revised Cochrane risk-of-bias tool for randomized trials (RoB 2), focused on the ROP outcomes relevant to this meta-analysis. Two reviewers (US and AKN) rated the trial independently and resolved any disagreements by consensus; the Henriksen *et al*. trial was judged to be at low risk of bias in all five domains and overall.

### Outcomes

Three ROP endpoints were analyzed, all as binary outcomes per infant: (i) any ROP, defined as ROP stage ≥1; (ii) ROP requiring treatment; and (iii) severe ROP, defined as stage 3 and/or treated.

### Statistical analysis

For each trial, gestational age-adjusted odds ratios (adjORs) with 95% confidence intervals (CIs) were either extracted from the published report or re-estimated from the raw data using logistic regression with treatment group and gestational age in completed weeks as predictors. Where standard maximum-likelihood estimation was unstable due to sparse events or complete/quasi-complete separation, models were refitted with Firth’s penalized likelihood, which provides separation-robust point estimates and profile-likelihood-based 95% CIs. Outcomes with zero events in both arms were classified as not estimable and excluded from pooling.

Crude risk ratios (RRs) from 2 × 2 contingency tables were pooled across trials using Mantel–Haenszel fixed-effect meta-analysis, matching the analytical strategy of Dang *et al*. Double-zero studies were excluded as uninformative on relative effect; a continuity correction of 0.5 was applied to studies with a single zero cell (default behavior of metabin).

Gestational age–adjusted ORs were pooled by inverse-variance random-effects meta-analysis. Between-study variance (τ^2^) was estimated by restricted maximum likelihood, and pooled CIs were derived using the Hartung–Knapp adjustment, which improves coverage when the number of studies is small. For studies whose adjOR was reported with an asymmetric (Firth or profile-likelihood) CI, the within-study standard error was approximated on the log scale as SE(log OR) = [log(CI_u_) − log(CI_l_)] / (2 × 1.96). This Wald-symmetry approximation is standard for aggregate-data meta-analysis; because the affected (sparse-event) studies carry small inverse-variance weight, the impact on the pooled estimate is negligible, although the within-study CI shown in the forest plot may differ marginally from the originally reported asymmetric CI. Between-study heterogeneity was quantified by Higgins’ I^2^ statistic and Cochran’s Q test. Pooled estimates are reported with 95% CIs. All analyses were performed in R version 4.3.2 (R Core Team, 2023) using the packages meta (v8.2-1), metafor, logistf (v1.26.x), readxl, dplyr, and tidyr.

## Results

### Any ROP

When all eligible trials contributed to the any-ROP analysis, the pooled estimate was consistent with that of Dang *et al*.: crude RR 0.87 [95% CI 0.71–1.08]; gestational age– adjusted OR 0.70 [0.46–1.08] (Figure 1a, d).

**Figure 1.**
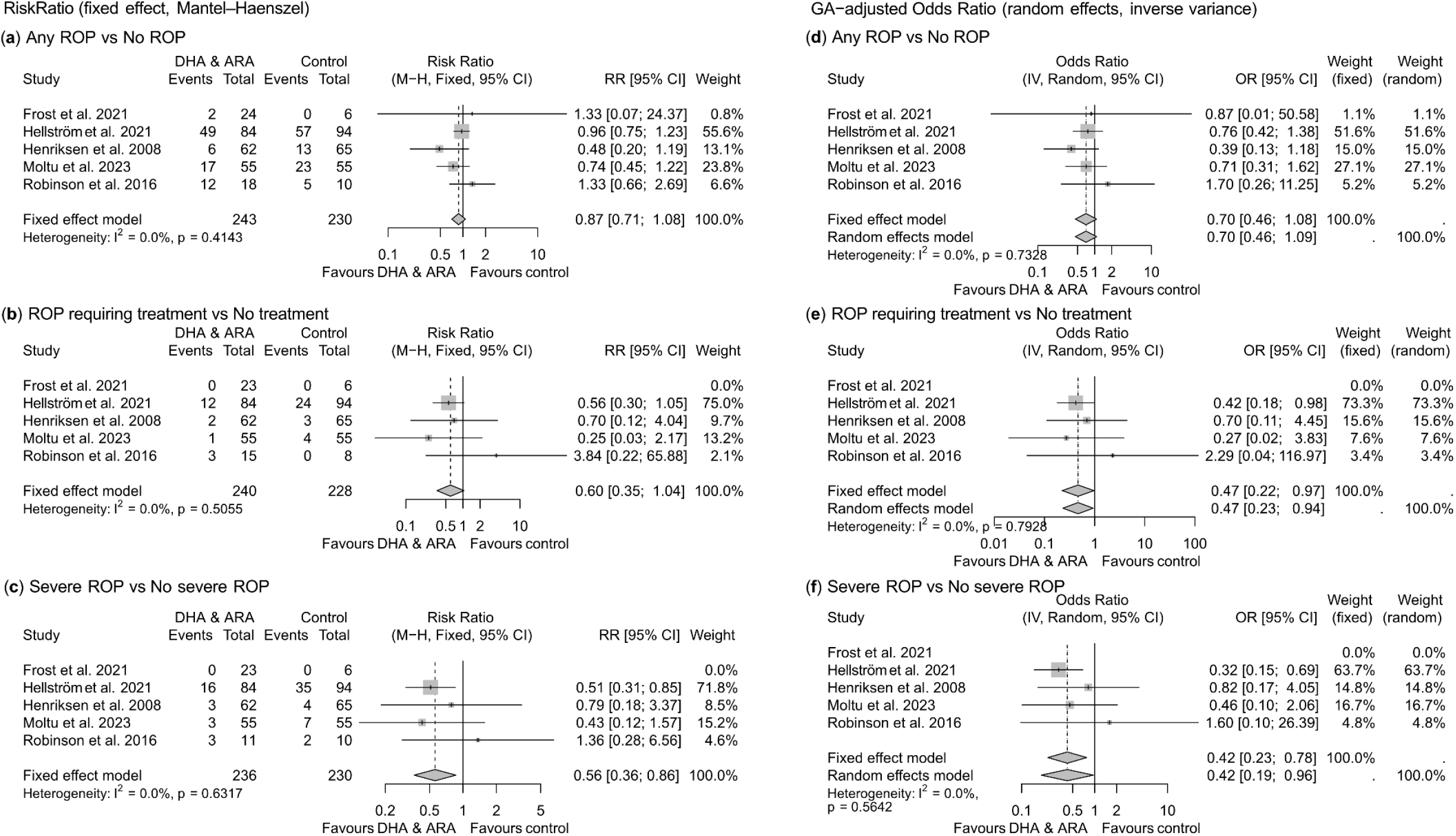
Meta-analyses of retinopathy of prematurity (ROP) outcomes following enteral supplementation with docosahexaenoic acid (DHA) and arachidonic acid (ARA). Panels **a–c** show pooled risk ratios (RR) from raw event counts using Mantel– Haenszel fixed-effect meta-analysis. Panels **d–f** show pooled gestational age–adjusted odds ratios (OR) using inverse-variance meta-analysis with restricted maximum likelihood (REML) τ^2^ estimation and Hartung–Knapp confidence-interval adjustment; for completeness, both fixed-effect and random-effects pooled estimates are reported. Outcomes are: (**a, d**) any ROP; (**b, e**) ROP requiring treatment; (**c, f**) severe ROP (stage 3 and/or treated). Within each panel, study identifiers, event counts and totals per arm (where applicable), and per-study effect estimates with 95% confidence intervals (CIs) are shown alongside the forest plot. Square area is proportional to study weight; horizontal lines represent 95% CIs; diamonds represent pooled estimates. For Robinson *et al*. 2016, the study protocol allowed determination of ROP status through discharge only; therefore, infants with uncertain outcomes (five infants with documented ROP for whom the final ROP stage was not reported, and two infants with stage 3 ROP for whom treatment status was not reported) were excluded from event counts. For Frost et al. 2021, one infant was lost to follow-up after ROP stage 1 or 2 had been documented and was excluded from event counts for severe and treated ROP.

### ROP requiring treatment

For ROP requiring treatment, the crude RR suggested a lower risk but did not reach statistical significance (RR 0.60 [0.35–1.04], Figure 1b), whereas the gestational age-adjusted OR indicated lower odds (adjOR 0.47 [0.23–0.94], Figure 1e) with enteral DHA+ARA supplementation.

### Severe ROP

For severe ROP, DHA+ARA supplementation produced a significant protective effect in both unadjusted and adjusted models: RR 0.56 [0.36–0.86]; adjOR 0.42 [0.19–0.96] (Figure 1c, f).

## Discussion

When this meta-analysis is repeated with complete outcome data from all eligible DHA+ARA trials and also including the clinically relevant endpoint severe ROP, the picture that emerges differs from that conveyed by Dang *et al*. Enteral DHA+ARA supplementation did not significantly reduce the occurrence of any ROP, consistent with the conclusion of Dang *et al*. for this endpoint, but significantly reduced the risk of severe ROP and, after adjustment for gestational age, also of ROP requiring treatment. Severe ROP carries a meaningful risk of life-long visual impairment, and any safe and inexpensive intervention that reduces its incidence has substantial clinical and societal value. Enteral DHA+ARA supplementation is logistically simple, low-cost, and already feasible in modern neonatal intensive care units, and the present re-analysis indicates that the available randomized evidence supports a protective effect on severe ROP. This preventive potential warrants explicit consideration in guideline deliberations and in the planning of future trials, which should be powered for severe ROP as the primary endpoint and should report ROP stage distributions and treatment status separately to enable complete pooled analysis.

### Strengths and limitations

Strengths of the present re-analysis include complete outcome records from every eligible trial, addition of the clinically relevant endpoint severe ROP, and the use of gestational age–adjusted odds ratios in addition to crude risk ratios.

The principal limitations follow from the underlying evidence base. Only five randomized controlled trials met the eligibility criteria, and DHA and ARA doses, supplementation duration, and concomitant nutritional practices vary across them. Distributions of gestational age and other risk factors also differ across trials, which we partly address by reporting gestational age-adjusted ORs.

## Conclusions

Including all eligible trials and adding severe ROP as an outcome, enteral DHA+ARA supplementation reduces the risk of severe ROP and, after adjustment for gestational age, also of ROP requiring treatment in preterm infants. These findings differ from those of Dang *et al*. and support reconsideration of DHA+ARA supplementation as a strategy to reduce sight-threatening ROP. The evidence base nonetheless remains limited; only five trials with heterogeneous dosing and a modest total enrolment contributed to the analysis, and adequately powered randomized trials with severe ROP as the primary outcome are warranted to confirm the magnitude of this effect.

## Declarations

### Author contributions

AKN and AH conceived the re-analysis; US and AP performed the statistical analyses; US, PL, AH contributed to methodology and interpretation; SJM, BF, DTR, CH contributed outcome data from their respective trials and to interpretation; all authors revised the manuscript critically for important intellectual content and approved the final version.

### Data availability

Pooled outcome data and analysis code are available from the corresponding author on reasonable request. Individual participant data from the included trials are subject to the respective trial data sharing agreements.

### Ethics

This work is a re-analysis of de-identified summary outcome data from previously published randomized controlled trials, each of which had ethics approval at its respective site. No new ethics approval was required for the present re-analysis.

### AI use

The authors used Claude (Anthropic) to refine language and improve clarity of the manuscript text. The AI was not used to generate scientific content, perform statistical analyses, or interpret results. All AI-suggested edits were reviewed and approved by the authors, who take full responsibility for the content of the submission.

## References

1. Dang, D., et al., Effect of enteral supplementation of DHA with or without ARA in preterm infants: a meta-analysis. Arch Dis Child Fetal Neonatal Ed, 2025. 111(1): p. F27–f33.

2. Hellström, A., L.E. Smith, and O. Dammann, Retinopathy of prematurity. Lancet, 2013. 382(9902): p. 1445–57.

3. Hellström, A., et al., Nutritional interventions to prevent retinopathy of prematurity. Pediatric Research, 2024. 96(4): p. 905–911.

4. Stoltz Sjöström, E., et al., Low energy intake during the first 4 weeks of life increases the risk for severe retinopathy of prematurity in extremely preterm infants. Arch Dis Child Fetal Neonatal Ed, 2016. 101(2): p. F108–13.

5. VanderVeen, D.K., et al., Early nutrition and weight gain in preterm newborns and the risk of retinopathy of prematurity. PLoS One, 2013. 8(5): p. e64325.

6. Martin, C.R., et al., Decreased postnatal docosahexaenoic and arachidonic acid blood levels in premature infants are associated with neonatal morbidities. The Journal of pediatrics, 2011. 159(5): p. 743–749.e7492.

7. Dyall, S.C., et al., Omega-3 and omega-6 polyunsaturated fatty acids in neurodevelopment and prematurity: Correcting imbalances and closing the Preterm PUFA Gap. Prog Lipid Res, 2026. 101: p. 101379.

8. Lapillonne, A., S. Eleni dit Trolli, and E. Kermorvant-Duchemin, Postnatal docosahexaenoic acid deficiency is an inevitable consequence of current recommendations and practice in preterm infants. Neonatology, 2010. 98(4): p. 397–403.

9. Boeck, M., et al., Combined dietary omega-3 and omega-6 fatty acids protect against hyperglycemia-associated retinopathy in neonatal mice. Pharmacol Res, 2025. 219: p. 107877.

10. Fu, Z., et al., Omega-3/Omega-6 Long-Chain Fatty Acid Imbalance in Phase I Retinopathy of Prematurity. Nutrients, 2022. 14(7).

11. Diggikar, S., et al., Effect of Enteral Long-Chain Polyunsaturated Fatty Acids on Retinopathy of Prematurity: A Systematic Review and Meta-Analysis. Neonatology, 2022. 119(5): p. 547–557.

12. Hellström, A., et al., Effect of Enteral Lipid Supplement on Severe Retinopathy of Prematurity: A Randomized Clinical Trial. JAMA Pediatr, 2021. 175(4): p. 359–367.

13. Moltu, S.J., et al., Arachidonic and docosahexaenoic acid supplementation and brain maturation in preterm infants; a double blind RCT. Clin Nutr, 2024. 43(1): p. 176–186.

14. Frost, B.L., et al., Randomized Controlled Trial of Early Docosahexaenoic Acid and Arachidonic Acid Enteral Supplementation in Very Low Birth Weight Infants. J Pediatr, 2021. 232: p. 23–30.e1.

15. Robinson, D.T., et al., Early docosahexaenoic and arachidonic acid supplementation in extremely-low-birth-weight infants. Pediatr Res, 2016. 80(4): p. 505–10.

16. Gao, Z., et al. Effect of enteral supplementation of DHA with or without ARA in preterm infants: A systematic review and meta-analysis. 2024 Cited 2026; Available from: https://www.crd.york.ac.uk/PROSPERO/view/CRD42024552578.

17. Henriksen, C., et al., Improved Cognitive Development Among Preterm Infants Attributable to Early Supplementation of Human Milk With Docosahexaenoic Acid and Arachidonic Acid. Pediatrics, 2008. 121(6): p. 1137–1145.

